# Development and Initial Validation of the Quality of life Evaluation in *NF2*-related Schwannomatosis Trials (QUEST) Assessment

**DOI:** 10.64898/2026.06.09.26355287

**Authors:** Vanessa L. Merker, Sophia C. Carias, Rosalie E. Ferner, John F. Golding, Scott R. Plotkin, Frank D. Buono

**Affiliations:** Department of Neurology and Mass General Brigham Cancer Institute, Massachusetts General Hospital, Boston, MA, USA; Department of Neurology, Guy’s and St. Thomas’ NHS Foundation Trust, London, UK; Department of Psychology, University of Westminster, London, UK; Department of Psychiatry, Yale School of Medicine, New Haven, CT, USA

**Keywords:** neurofibromatosis 2, quality of life, patient reported outcome measure, psychometrics, clinical trials

## Abstract

Individuals with *NF2*-related schwannomatosis (*NF2*-SWN) experience a complex constellation of physical, emotional, and social symptoms that substantially impact quality of life (QoL). Although disease-specific patient-reported outcome measures are increasingly important for evaluating treatment benefit in clinical trials, existing *NF2*-SWN QoL measures have limitations in content coverage and sensitivity to change. This study describes the development and initial validation a new disease-specific QoL assessment - the <u>Qu</u>ality of Life <u>E</u>valuation in *NF2*-related <u>S</u>chwannomatosis <u>T</u>rials (QUEST). Using a three-phase, mixed-methods approach, items were generated through concept elicitation interviews with individuals with *NF2*-SWN and clinicians, prioritized via patient survey data, and refined through iterative cognitive debriefing procedures. The resulting 21-item QUEST assesses the extent to which *NF2*-SWN has negatively impacted a person’s daily life over the past seven days. Initial psychometric evaluation was conducted in an international sample of 174 individuals with *NF2*-SWN aged 15 years and older (117 women (67%), 158 White individuals (89%)). Exploratory factor analysis supported a four-factor structure, and the total score demonstrated excellent internal consistency and strong test–retest reliability. Evidence of construct validity was demonstrated through hypothesized associations with disease-specific, generic, and domain-specific QoL measures, as well as known-groups validity based on self-reported disease severity and number of prior surgeries. Incremental validity analyses indicated that QUEST explained unique variance beyond existing measures. Together, findings support the QUEST as a reliable and valid disease-specific QoL measure with strong content validity and feasibility for use as a clinical trial endpoint in *NF2*-SWN.

**Public Significance Statement:** People with *NF2*-related schwannomatosis experience symptoms that can affect many areas of daily life, yet these impacts are often difficult to measure in clinical trials. We developed a new questionnaire with direct patient input to capture how the condition affects quality of life, which can help researchers and clinicians better evaluate whether treatments improve outcomes that matter to patients.

## Background

*NF2*-related schwannomatosis (*NF2*-SWN) is a rare, neurogenetic disorder that predisposes individuals to develop bilateral vestibular schwannomas as well as other tumors in the brain, spine, and peripheral nerves. *NF2*-SWN has a prevalence of approximately 1 in 58,000, with approximately 25-50% of individuals inheriting the disorder from a parent (Forde et al., 2024). Individuals are most commonly diagnosed in their 20s (Evans et al., 1992), although affected individuals may show symptoms as early as birth (Ruggieri et al., 2016). The majority of *NF2*-SWN patients become deaf over their lifetime, and patients may experience a range of additional symptoms including tinnitus, balance problems, facial weakness, pain, difficulty walking, and ophthalmologic issues including juvenile cataracts and retinal abnormalities (Plotkin & Wick, 2018). Collectively, these symptoms can greatly impact quality of life (QoL), including impacts on physical functioning, activities of daily living, life roles, social integration, and emotional wellbeing (Carias et al., 2025; Merker et al., 2016).

Measuring QoL in people with *NF2*-SWN is important to assess the effectiveness of routine clinical care options as well as novel therapeutics within clinical trials (Slade et al., 2018). Quality of life and other patient-reported outcomes (PROs) have particularly grown in importance to regulatory bodies and health technology assessment agencies as they strive to consider patient-reported clinical benefit during drug approval, labeling, and reimbursement decisions (Doward et al., 2010; Food and Drug Administration, 2009; Kieffer et al., 2020; Pe et al., 2025). For example, the Food and Drug Administration (FDA) launched a significant effort to incorporate the patient voice in drug development (Food and Drug Administration, 2020, 2022a), including using PROs to capture the impacts of treatment that matter most to patients (Food and Drug Administration, 2022b, 2023). As disease-specific measures may be more specific to patient concerns and sensitive to health status change than generic measures (Patrick et al., 2007), disease-specific QoL measures are critical to understanding the benefits and drawbacks of new therapies on *NF2*-SWN patients’ physical, mental, and social wellbeing.

However, currently available measures of disease-specific QoL in *NF2*-SWN have several drawbacks for use in clinical trials. The Neurofibromatosis Two Impact on Quality of Life scale (NFTI-QoL)(Hornigold et al., 2012) is the most commonly used measure in clinical care (Lawson McLean et al., 2025) and is currently recommended for use in clinical trials by the Response Evaluation in Neurofibromatosis and Schwannomatosis (REiNS) International Collaboration (Wolters et al., 2021). However, the REiNS Patient Reported Outcomes (PRO) working group noted key potential limitations in the measure, including that the 8 items may not assess all important *NF2*-SWN symptoms, and questionnaire features may limit sensitivity to change within clinical trials. A prior study using the NFTI-QoL observed ceiling effects in multiple items (Hamoy-Jimenez et al., 2020), and cognitive debriefing interviews with participants in a recent trial (Plotkin et al., 2024) confirmed several instances where the measure did not capture meaningful QoL improvements (Von Imhof et al., 2023), leading to further concern about the responsiveness of this measure as a clinical trial endpoint. Other existing measures of disease-specific quality of life for *NF2*-SWN patients were also found to have major limitations by REINS (Wolters et al., 2021), including lack of adequate psychometric validation, limited feasibility, or lack of comprehensiveness due to an exclusive focus on one tumor type. Together, these studies suggest that developing and validating a new disease-specific QoL measure according to current best practice guidance for patient reported outcomes (Mokkink et al., 2010; Patrick et al., 2011a, 2011b) would be a valuable addition to *NF2*-SWN clinical trial design.

The present study used a three-phase, mixed-methods approach to develop and test the initial psychometric properties of the <u>Qu</u>ality of life <u>E</u>valuation in *NF2*-related <u>S</u>chwannomatosis <u>T</u>rials (QUEST) assessment, a new patient-reported QoL measure designed to assess how much *NF2*-SWN symptoms and issues have negatively impacted an individual over the past 7 days. This focus was selected to ensure the measure would be suitable for use in clinical trials testing new treatments and interventions for *NF2*-SWN patients. In Phase I, we conducted concept elicitation interviews with 16 *NF2*-SWN patients and 10 *NF2*-SWN clinicians and surveyed 58 *NF2*-SWN patients to generate items and prioritize them for inclusion in the assessment. In Phase II, we conducted 4 iterative rounds of cognitive debriefing with an additional 16 *NF2*-SWN patients and consulted with experts in NF patient-reported outcomes to finalize the assessment wording and format. In Phase III, the 22-item QUEST measure was administered to 174 *NF2*-SWN patients to assess its factor structure, reliability, and validity.

### Transparency and Openness

This study’s design and its analysis were not preregistered. Data for Phase I of the research is not publicly available; data for Phase II and Phase III will be publicly available no later than October 1, 2027 at the NF Data Portal and can be accessed at https://nf.synapse.org/Explore/Studies/syn53181385/. Analysis code is not available for the study; however, other research materials are available on request from the first author. Analyses were performed in NVivo Version 1.7.2 (Lumivero, 2022); Excel for Microsoft 365 (Microsoft, 2024); and SPSS, version 31 (IBM, 2025).

## Phase I: Item Generation and Item Selection

The aim of this phase of research was to 1) generate a comprehensive list of potentially relevant items through concept elicitation interviews with patients and clinicians and 2) prioritize items for inclusion in the final assessment using a combination of statistical analysis, survey input from additional patients, and team discussion. These procedures were undertaken to ensure our assessment’s content validity (specifically, comprehensiveness and relevance to our target population). Study procedures were reviewed and approved by the Dana Farber/Harvard Cancer Center Institutional Review Board (Protocol 19-828) or the Mass General Brigham Institutional Review Board (Protocol 2023P003305), and no remuneration was provided to participants in this phase of research.

### Phase 1: Method

#### Interview Participants

Patients were recruited from stage 1 of the brigatinib treatment arm of the INTUITT-NF2 trial (NCT04374305), an adaptive platform-basket trial testing the efficacy of multiple chemotherapies in treating nervous system tumors in people with *NF2*-SWN. Per the trial’s inclusion criteria, all participants were ≥12 years, had a clinical diagnosis of *NF2*-SWN, and had at least one *NF2*-related tumor (i.e., vestibular schwannoma, non-vestibular schwannoma, meningiomas, or ependymoma) with documented radiographic progression over the prior 3 years (full inclusion and exclusion criteria available at (Plotkin et al., 2024)). This sample was selected to ensure generated items reflected the impact of all tumor types characteristic of *NF2*-SWN, including the potentially more wide-ranging or severe symptoms of individuals with progressive disease.

Clinicians were eligible for the study if they practiced in the United States, United Kingdom, or Australia and had self-identified expertise in caring for patients with *NF2*-SWN and/or experience as an investigator on clinical trials for *NF2*-SWN. Potentially eligible clinicians were identified through their publications in PubMed, postings in ClinicalTrials.gov, or membership in the *NF2*-SWN committee of the Neurofibromatosis Clinical Trial Consortium. Study researchers purposively sampled clinicians across a range of medical specialties relevant to *NF2*-SWN and sequentially contacted individuals by email until ten agreed to participate.

#### Survey Participants

Individuals with self-reported *NF2*-SWN ages ≥12 years who lived in the United States and were willing and able to complete an online survey were eligible for this portion of the study. Individuals were recruited from the Children’s Tumor Foundation NF Registry (Seidlin et al., 2017). Registry staff emailed a flyer describing the study to all NF Registry participants who met the above criteria and had consented to be contacted about future research opportunities.

#### Interview Procedure

Interview procedure is described in detail in (Carias et al., 2025). In brief, *NF2*-SWN patients were consented to the study by their local treating physicians across four U.S. academic medical centers and then invited to schedule interviews with the centralized staff overseeing qualitative procedures. Patients completed semi-structured qualitative interviews using videoconferencing software after approximately 6 months or 12 months on treatment, or at the time they stopped treatment. To facilitate interviews for individuals with hearing loss, use of communication access real-time transcription (CART) or American Sign Language (ASL) interpreters was offered to all participants. Interviews included open-ended questions about the pre-treatment impacts of *NF2*-SWN on their QoL, followed by a request to comment on whether they had experience any of the *NF2*-SWN related issues included in the NFTI-QoL (Hornigold et al., 2012). Similarly, clinicians completed semi-structured video interviews after giving verbal consent to participate. Interview questions focused on assessing the impact of *NF2*-SWN on patients’ quality of life across physical, emotional, and social domains. All patient and clinician interviews were audio-recorded, transcribed verbatim, de-identified, and reviewed by a research assistant for accuracy.

#### Survey Procedure

Recruitment flyers directed interested participants to access a REDCap survey (Harris et al., 2009), where they were shown a consent fact sheet describing the study and asked questions to confirm their eligibility. Individuals who met eligibility criteria and checked a box indicating that they wished to participate in the study then proceeded directly to the research survey. Participants were asked to provide demographic and clinical information (age, gender, race, ethnicity, whether either of their parents had *NF2*-SWN, their self-reported symptom severity, and whether they had ever been in an *NF2*-SWN clinical trial). Finally, participants were asked to rate the importance of various concepts identified from the patient and clinician interviews. Specifically, participants were asked “How important are the following to your quality of life regarding *NF2*-SWN?”, shown the list of concepts, and asked to select a response for each using a 5-point scale (1=not at all, 2=slightly, 3=somewhat, 4=moderately, or 5=extremely important).

#### Data analysis

For patient interviews, a hybrid inductive and deductive coding process was completed using NVivo software by two independent raters. Deductive codes were derived from concepts already included in the NFTI-QoL (Hornigold et al., 2012); inductive codes were created for new concepts raised by participants and iteratively added to the codebook based on consensus discussion between raters. After completing this process for all patient interviews, the final patient codebook was deductively applied to clinician interview transcripts directly in Microsoft Word by a single rater; new concepts raised only by clinicians were added as needed. Concepts were recorded according to the level of detail provided by interviewees; for this reason, some symptoms are captured at both an overall level (e.g. pain) and with more specificity when such detail was provided (e.g. intensity of pain, frequency of pain, etc.).

After the number of patients and clinicians endorsing each concept was tallied, a scale-level content validity index (CVI-S) procedure was used to identify what proportion of concepts had sufficient agreement amongst patients and/or clinicians. The content validity index (CVI) is the mostly widely accepted and recognized assessment for evaluating rating scales for content validity (Lawshe, 1975; Lynn, 1986). The CVI-S formula minimizes inherent human error and provides a more reliable assessment of the value of items for inclusion in an overall measure. The CVI-S threshold was measured independently for patient interviews and clinician interviews, and all concepts within at least two standard deviations of the CVI-S threshold were included in the subsequent patient survey. For the survey, all demographics are summarized using descriptive statistics; patient ratings on the importance of each concept are described by the percentage of participants selecting each response option and the group-level mean and standard deviation of the Likert scale ratings (on a 1-5 scale).

### Phase 1: Results

We conducted concept elicitation interviews with 16 individuals with *NF2*-SWN and 10 *NF2*-SWN clinicians. Patient with *NF2*-SWN ranged in age from 15-54 years (median, 25 years); 11 (69%) were female and 5 (31%) were male. The majority were white (14, 88%) with 1 patient (6%) each identifying as Asian or as other race (not including black, Native American, or Native Hawaiian/Pacific Islander). Two patients (13%) were Hispanic/Latino, with the majority non-Hispanic/Latino (12, 75%) or unknown (2, 13%). Clinicians were half female and half male (n=5 each), and race/ethnicity was not collected for these participants. They included 3 neuro-oncologists, 2 neurologists, 2 surgeons, 1 psychologist, and 1 advance practice nursing provider.

Analysis of concept elicitation interviews yielded 83 symptoms or QoL impacts of *NF2*-SWN [Supplemental Table 1]. This included general statements on symptom areas (e.g. pain); descriptions of specific aspects of symptoms that impacted quality of life (e.g. frequency of pain, intensity of pain, and duration of pain), and aspects of quality of life impacted by one or more *NF2*-SWN symptoms (e.g., physical fatigue, ability to engage in hobbies and leisure activities, feeling a lack of control). Of the 83 concepts, 48 were mentioned by patients and clinicians, 20 were mentioned only by patients, and 15 were mentioned only by clinicians.

CVI-S analysis indicated that 17 items had consensus among patients or clinicians, and all 35 concepts within at least two standard deviations of the CVI-S threshold (i.e., had a CVI-S ≥0.4, equivalent to an endorsement from 7/16 patients and/or 4/10 clinicians) were advanced for further evaluation in a national survey of patients with *NF2*-SWN. 58 individuals with *NF2*-SWN rated the importance of each concept to their own quality of life. The median age of participants was 45 years old (range, 15-85 years); additional self-reported demographic and clinical characteristics of survey participants are shown in Table 1.

**Table 1.** Self-reported demographic and clinical characteristics of Phase 1 survey participants.

|  | N (%) |
| --- | --- |
| Gender |  |
| Female | 39 (67.2%) |
| Male | 19 (32.8%) |
| Non-binary or none of the above | 0 (0.0%) |
| Race<sup>1</sup> |  |
| White | 56 (96.6%) |
| Black or of African Ancestry | 1 (1.7%) |
| American Indian, Alaska Native, or Other Indigenous | 1 (1.7%) |
| Asian | 0 (0.0%) |
| Native Hawaiian or Pacific Islander | 0 (0.0%) |
| Other | 1 (1.7%) |
| Ethnicity |  |
| Hispanic/Latino/Spanish origin | 5 (8.6%) |
| Non-Hispanic/Latino/Spanish origin | 53 (91.4%) |
| Familial *NF2*-SWN |  |
| Yes | 9 (15.5%) |
| No | 48 (82.8%) |
| Unsure | 1 (0.0%) |
| Self-reported *NF2*-SWN Severity |  |
| No symptoms | 0 (0.0%) |
| Mild: symptoms do not usually affect daily life | 10 (17.2%) |
| Moderate: symptoms affect daily life but are not severe | 36 (62.1%) |
| Severe: symptoms significantly impact daily life | 12 (20.7%) |
| Previous Participation in Clinical Trial |  |
| Yes | 17 (29.3%) |
| No | 35 (60.3%) |
| Unsure | 6 (10.3) |

<sup>1</sup>Participants could select more than one option, so totals sum to more than 100%.

Participants’ ratings of the importance of each item to their quality of life are shown in Table 2. The items with the highest mean ratings were hearing, balance, quality of relationships, vision, and mobility (which were rated as “extremely important” by 74.5%, 56.4%, 58.6%, 67.9%, and 61.8% of respondents, respectively.) The items with the lowest mean ratings showed a bimodal distribution with relatively high rates of individuals indicating this was “not at all important” to them (51.8% for worry about having kids, 25.5% for difficulty carrying out work/school-related tasks, and 25.5% for ability to care for personal hygiene and dressing.) These results suggest that family planning and employment status may be highly salient for some individuals with NF2-SWN at particular life stages, but are not universally relevant across the broader population. Many respondents may have already completed family building, chosen not to have children, or may not currently be working or enrolled in school.

**Table 2.**
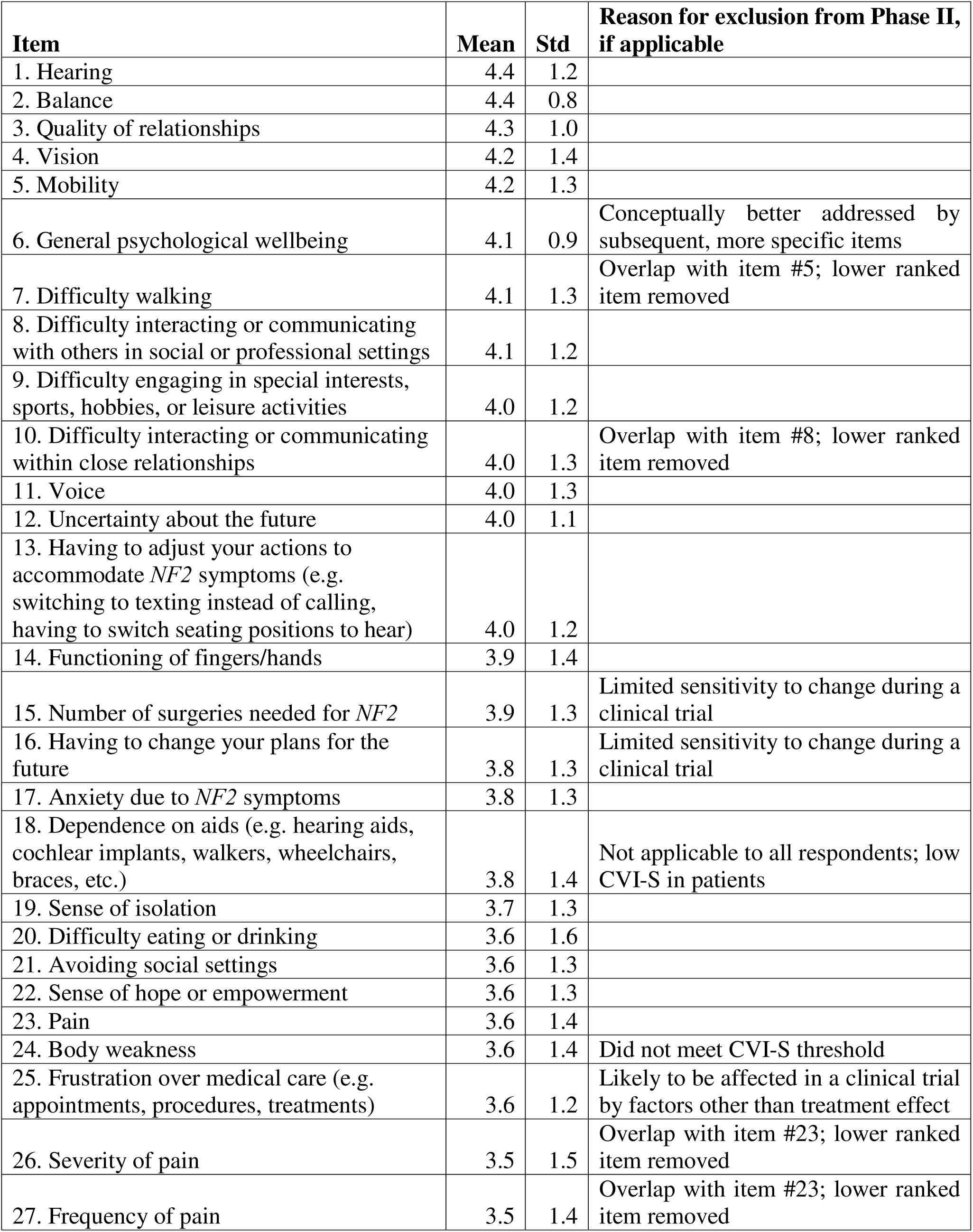
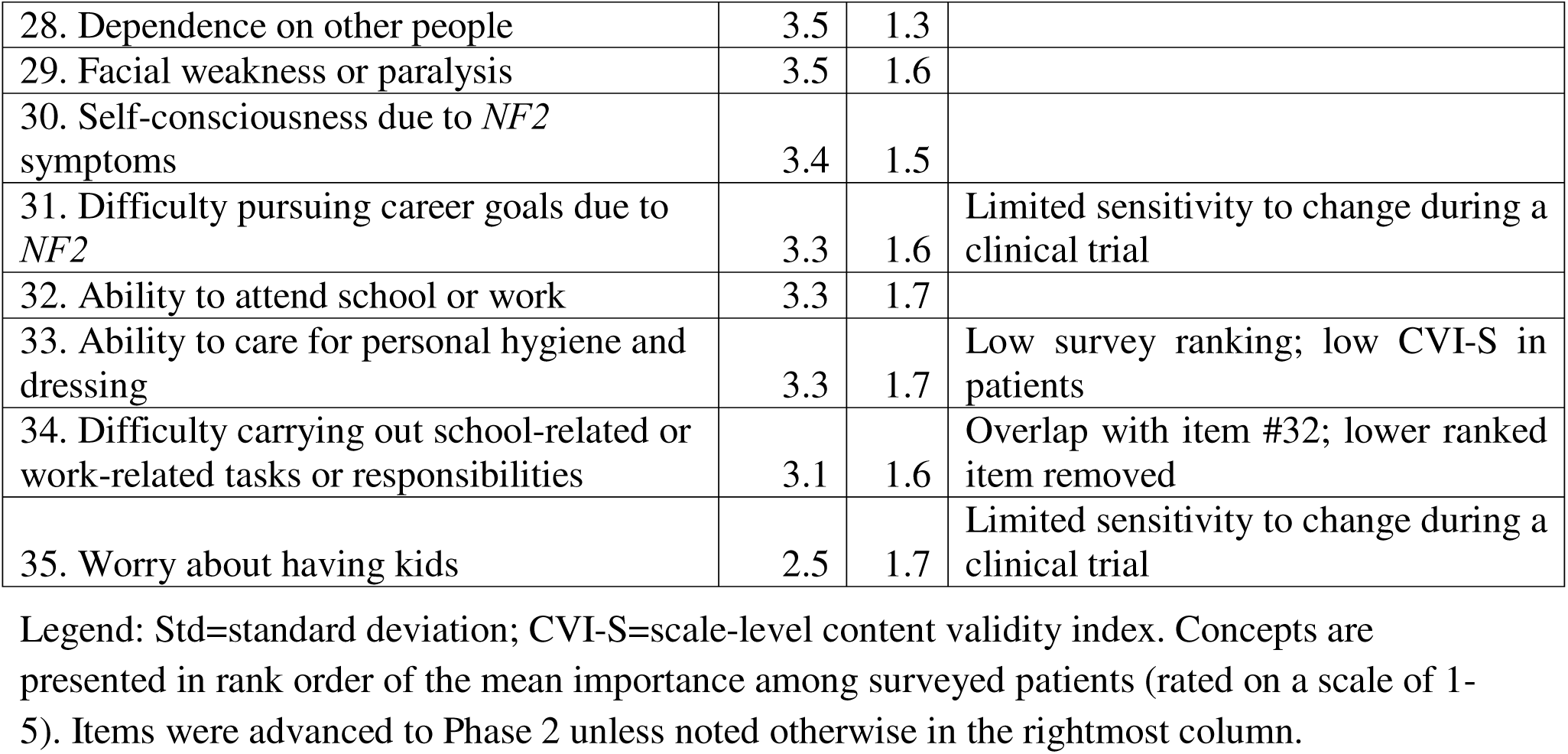
Patient Ratings of the Importance of Concepts to their Quality of Life.

Ultimately, 21 of the 35 concepts were selected for the draft measure based on CVI-S results, survey ratings, and practical considerations such as suitability for use in clinical trials. As *NF2*-SWN clinical trials typically evaluate treatment effects over 1-2 years, we excluded concepts unlikely change meaningfully over this timeframe (e.g. long-term future plans, career goals, and family-planning decisions). Although the number of surgeries is an important correlate of quality of life in NF2-SWN, we also excluded this concept because surgery frequency is unlikely to increase during a relatively short trial period; moreover, undergoing surgery during a trial may prompt treatment interruption or discontinuation. Finally, frustration with medical care was excluded because it may be influenced by non-treatment related factors during a clinical trial (e.g., additional research appointments and assessments), reducing the measure’s utility as a clinical trial outcome endpoint.

## Phase II: Cognitive Debriefing and Measure Development

The aims of this phase of research were to 1) iteratively refine measure wording to ensure participants could adequately understand and respond to items, and 2) optimize the electronic format to enhance participant experience and maximize assessment feasibility. These procedures were undertaken to ensure comprehensibility and content validity of the assessment within its intended patient population and context of use. Study procedures were reviewed and approved by the Mass General Brigham Institutional Review Board (Protocol 2023P003305).

### Participants

Individuals with *NF2*-SWN ages ≥12 years living in the United States who were willing and able to complete a recorded, virtual interview were eligible for the study. Participants were recruited from the Massachusetts General Hospital NF Clinic, the Children’s Tumor Foundation NF Patient Registry, and other NF patient advocacy organizations across the U.S. who distributed study flyers in their newsletters or private social media channels. Individuals were purposively sampled based on age, gender, and self-reported disease severity to ensure broad understanding and applicability of item content.

### Procedure

After providing verbal consent to participate in the study, participants provided demographic information and scheduled a one-on-one, virtual interview with a member of the research team trained in qualitative methods that lasted between 60 and 90 minutes. CART was offered to all participants with hearing loss. Interviews followed a retrospective debriefing protocol, wherein participants first accessed an online REDCAP survey to complete the draft QUEST measure and seven survey items assessing the measure’s content validity (Brod et al., 2009). Participants then immediately completed a semi-structured interview that included questions about how they understood the instructions, each item, the response options, and recall period, as well as any other feedback on the measure or suggestions for improvement. Following interview completion, a $25 Amazon gift card was provided as remuneration. All interviews were audio-recorded and transcribed verbatim.

### Measures

#### Quality of life Evaluation in NF2-related Schwannomatosis Trials (QUEST)

The initial draft measure contained the 21 concepts identified in Phase 1 as well as one summary item asking participants about their *NF2*-SWN symptoms overall. Concepts related to *NF2*-SWN symptoms were phrased to identify the symptom impact on people’s daily activities (e.g. “How much has *NF2*-related pain interfered with your day-to-day activities?”) and concepts that identified a larger quality of life impact were phrased to isolate the impact of *NF2*-symptoms on each domain (e.g., “How much have *NF2*-related symptoms made you feel dependent on others?”) A 5-point response scale was chosen to provide adequate discrimination but avoid an overwhelming number of options. All items were negatively worded to simplify scoring. Of note, since the nomenclature for *NF2*-SWN has only recently changed from neurofibromatosis type 2 (*NF2*) to *NF2*-related schwannomatosis (Plotkin et al., 2022), we used the most common abbreviation used by patients (*NF2*) throughout the measure and explained this abbreviation in the instructions. Instructions also asked individuals to rate how much *NF2*-SWN symptoms had impacted them in the past 7 days; this recall period was chosen to reflect length of time that participants could accurately recall and to align with other NF clinical trial measures.

#### Content Validity Index

Interviewees were asked to rate the QUEST in terms of clarity and relevancy to the construct on a 4-point ordinal scale (strongly disagree, disagree, agree, strongly agree) across seven questions: 1) provided enough detail about how *NF2*-SWN impacts their QoL, 2) had confusing questions, 3) focused only on the aspects of QoL related to their *NF2*-SWN, 4) was too long, 5) would be helpful for patients with *NF2*-SWN, 6) would be helpful for doctors who treat patients with *NF2*-SWN, and 7) will sufficiently track QoL in *NF2*-SWN patients over time. To obtain a CVI rating, the number of participants (content experts) judging the item as relevant or clear (rating 3 or 4, reverse scored as appropriate) was divided by the number of participants providing a rating.

### Analysis

Rapid qualitative analysis was used to review participant feedback in real-time to make iterative adjustments to the draft measure prior to subsequent interviews (Gale et al., 2013; Gale et al., 2019). A single researcher summarized participant comments into an Excel spreadsheet, in which each participant was a single row, and each column denoted the area of the QUEST for which feedback was provided (i.e. instructions, recall period, response scale, each individual item, and any additional feedback). During analysis, two additional columns were added to reflect feedback applicable to multiple items (preferred wording to describe “daily activities” and “interference” of *NF2*-SWN symptoms with these activities).

We conducted four rounds of cognitive debriefing interviews with four participants per round to align with COSMIN guidelines for ensuring content validity during PRO development. Participant feedback was reviewed by three researchers (VM, SC, FB) after each round of interviews, with reference back to participant transcripts and quotes as needed. Based on consensus discussion, iterative changes were made to improve patient comprehension of items as intended and improve the measure’s ease of use in an electronic format. In addition, we consulted the Response Evaluation in Neurofibromatosis and Schwannomatosis (REiNS) International Collaboration Patient Reported Outcomes working group in between rounds 2 and 3 to seek feedback on the questionnaire as a whole, as well as appropriate interpretation of participant feedback regarding response options and the recall period, and again after the final round to confirm an appropriate age range for the measure. The final QUEST measure was created based on this input, in consultation with two additional research team members (RF, JG) experienced in *NF2*-SWN and questionnaire design.

### Phase II Results

Sixteen people with *NF2*-SWN participated in interviews. Participants had a median age of 31 years (range, 12-58); 50% were male and 50% were female; 13 (81%) were white, 1 (6%) was Asian, 1 (6%) was Black, and 1 (6%) declined to provide a race; and three participants (19%) had familial *NF2*-SWN. Disease severity was self-reported as no symptoms (n=1, 6%), mild (n=3, 19%), moderate (n=10, 63%), and severe (n=2, 13%).

The draft QUEST measure was refined based on analysis of interviews. This included changing the wording of items (e.g., changing “accommodations” to “adjustments”) or adding clarifications (e.g., that ratings of the impact of hearing loss should take into account any hearing devices participants use) to ensure item meaning was clear to participants of different ages and countries of residence. Based on our youngest participants’ difficulty understanding multiple concepts and discussion with REINS members, the recommended age range for this measure was determined to be ages 15 and up. Labels for the response scale were changed to ensure consistent interpretation, ultimately ranging from 1=Not at all to 5=Very Much (with 3=Somewhat, and options 2 and 4 unlabeled). The measure instructions were edited, with certain words bolded or underlined for emphasis based on participants’ suggestions. Finally, formatting was modified to ensure usability across electronic devices, including appearance on smaller phone screens and the addition of accessibility features such as modifiable text size and audio buttons so that content would be read aloud.

The gold-standard in evaluating content is the item-level context validity index (I-CVI) (Zamanzadeh et al., 2015). For the I-CVI the number of items considered relevant by all the judges is divided by the total number of items. In the average approach, the sum of I-CVIs is divided by the total number of items (Polit et al., 2007). Across the seven-item questionnaire, and the number of reviewers, the total score was .83. Acceptable levels for new assessments based on the criterion established is .80 (Abdollahpour et al., 2010).

## Phase III: Initial Psychometric Validation of Assessment

The aim of this phase of research was to determine the initial psychometric properties of the new assessment, including examining factor structure, reliability, and construct validity. Study procedures were reviewed and approved by the Mass General Brigham Institutional Review Board (Protocol 2024P003196).

### Phase III Method

#### Participants

Individuals with *NF2*-SWN ages ≥15 years from the United States, Canada, United Kingdom, and Australia were recruited from the Massachusetts General Hospital NF Clinic, other NF clinics (including INTUITT-NF2 trial sites), the Children’s Tumor Foundation NF Patient Registry, and multiple NF patient advocacy organizations. Recruitment occurred from January 2025 to June 2025.

#### Procedure

Participants completed an online survey in REDCap, including self-reported demographic and clinical data and several patient-reported outcome measures related to quality of life. Participants could optionally retake the QUEST measure two weeks later to assess test-retest reliability; individuals opting in were sequentially invited to do so until at least 80 responses were received. Participants received $20 remuneration (or local currency equivalent) for the initial survey and an additional $10 for the retake in the form of a Tango gift card. Fraudulent responses and bots were excluded from the dataset based on a combination of factors including the time to complete the survey being too quick, suspicious email addresses, repeated names, non-matching ages and dates of birth, completion of a hidden email field not visible to participants, inaccurate verification of submitted information in follow-up emails, and/or survey submission during a “bot attack” (a short period of time when hundreds of responses were submitted).

#### Measures

##### Quality of Life Evaluation in NF2-Related Schwannomatosis Trials

The final QUEST measure derived from Phase II of the study was used for all participants. A paper copy is provided for reference in Appendix A; the measure formatted for electronic administration and scoring manual are freely available upon request to the corresponding author. The QUEST total score is based on responses to 21 items and ranges from 21-105 with higher scores indicating worse quality of life; an additional, final item assessing the overall impact of *NF2*-related symptoms on participants day-to day activities on a scale of 0-10 was analyzed separately as a measure of global severity.

##### Neurofibromatosis 2 Impact on Quality of Life

The NFTI-QoL is an 8-item measure of disease-related quality of life for *NF2*-SWN patients (Hornigold et al., 2012) that was previously recommended for use in *NF2*-SWN clinical trials by the REiNS International Collaboration (Wolters et al., 2021). It showed high internal reliability (Cronbach’s alpha = 0.85) and test-retest reliability (r=0.84) within patients in the United Kingdom (Ferner et al., 2014)

##### Self-Assessment of Communication (SAC)

The SAC is a patient-reported measure of hearing function and hearing-related quality of life, which has been recommended for use in *NF2*-SWN clinical trials by the REiNS International Collaboration (Thompson et al., 2021). The 10-item SAC was administered to adults ages ≥18 years; the SAC-Adolescent (SAC-A) is a parallel 12-item measure that was administered to adolescents <18 years. Both versions have shown good internal consistency reliability (0.9 and 0.76, respectively) and test-retest reliability (0.8 and 0.76, respectively) (Hickson et al., 2007; Schow & Nerbonne, 1982; Wright et al., 2010).

##### Functional Assessment of Cancer Therapy – General, Version 4 (FACT-G)

The FACT-G assess general quality of life across four susbcales - physical, social/family, emotional, and functional wellbeing – and is recommended to assess generic (non-disease-specific) quality of life in trials testing medical products in adults with *NF2*-SWN by the REiNS International Collaboration (Wolters et al., 2021). All questions in the FACT-G use a 5-point rating scale (0 = Not at all; 1 = A little bit; 2 = Somewhat; 3 = Quite a bit; and 4 = Very much). The FACT-G total score is computed as the sum of the four subscale scores, provided the overall item response is at least 80% (i.e. at least 22 of the 27 items were answered) and has a possible range of 0-108 points, with higher scores indicating better quality of life. Cronbach’s alpha coefficients for the total scale typically ranges from 0.88 to 0.90 (Webster et al., 2003).

##### PROMIS Cognitive Function (PROMIS CF)

The PROMIS CF scale was administered as a computer adaptive test imported from the REDCap Shared Library, based on the Adult Bank v2.0 (for participants ≥18 years) or the Pediatric Bank v1.0 (for participants <18 years). A T-score of 50 indicates average levels of functioning as defined by the US calibration sample (standard deviation [SD] = 10). Cronbach’s alpha coefficients frequently reported between 0.94 and 0.97 in clinical and adult populations (Becker et al., 2014).

##### PROMIS Pain Interference (PROMIS PI)

The PROMIS PI was administered as a computer adaptive test imported from the REDCap Shared Library, based on the Adult Bank v1.1 (for participants ≥18 years) or the Pediatric Bank v2.0 (for participants <18 years). A T-score of 50 indicates average levels of functioning as defined by the US calibration sample (standard deviation [SD] = 10), which consists of health. PROMIS-PI is a psychometrically sound instrument with good reliability and Cronbach alpha of .88 (Amtmann et al., 2010).

#### Data analysis

We assessed the QUEST measure’s structural validity using exploratory factor analysis with varimax rotation, specifically a Kaiser-Meyer-Olkin (KMO) measure for sampling adequacy and Bartlett’s test of sphericity. We considered a KMO value of >0.5 and a p-value of ≤0.05 for Bartlett’s test to provide adequate evidence of the factor structure. We assessed the measure’s internal consistency reliability using Cronbach’s alpha and test-retest reliability using intraclass correlation coefficient, using a threshold of >0.70 to retain items in the final measure. To assess convergent validity, we hypothesized that there would be a large strength correlation (r≥0.70) of the QUEST to another disease-specific QoL measures (NFTI-QoL) and medium strength correlation (r≥0.40) of the QUEST to generic QoL (FACT-G) or domain-specific measures (SAC; PROMIS). Known-groups validity was assessed by testing for statistically significant differences (p<0.05) in scale scores between groups of participants with different patient-reported disease severity and number of surgeries using ANOVAs.

### Phase III: Results

#### Demographics

A total of 174 individuals participated in the research study (Table 3). Average age was 41.9 years (SD 16.3; range, 15-79 years). Most (67%) were female and white (89%), and half (50%) were working full-time. Just over half of participants had moderate disease severity with 1-4 prior surgeries due to their *NF2*-SWN. The average time it took participants to complete the QUEST measure on first administration was 4 minutes and 3 seconds (excluding three outliers who completed the questionnaire over >24 hours.) One hundred sixty participants agreed to be recontacted for an optional retake of the QUEST; of these, the first 107 were sequentially invited to participate in the retake until 81 (76%) completed their response.

**Table 3.** Phase 3 Survey Participant Demographics.

|  | **Frequency** | **Percentage** |
| --- | --- | --- |
| **Gender** |  |  |
| Male | 55 | 31% |
| Female | 117 | 67% |
| Non-Binary | 1 | 1% |
| None of these describe me | 1 | 1% |
| **Race** |  |  |
| White | 158 | 89% |
| Black | 6 | 3% |
| American Indian | 1 | 1% |
| Asian | 9 | 4% |
| Native Hawaiian/Pacific Islander | 0 | 0% |
| Other | 6 | 3% |
| **Employment Status** |  |  |
| Full Time | 89 | 50% |
| Temporarily on Leave | 8 | 3% |
| Unemployed but looking | 8 | 3% |
| Retired | 20 | 10% |
| Disabled | 36 | 19% |
| Homemaker | 11 | 5% |
| Student | 18 | 9% |
| Other | 1 | 0% |
| Prefer not say | 3 | 1% |
| Not sure | 0 | 0% |
| **Familial NF** |  |  |
| Yes | 46 | 26% |
| No | 116 | 67% |
| Unsure | 12 | 7% |
| **Self-reported Severity** |  |  |
| No symptoms | 3 | 2% |
| Mild | 35 | 21% |
| Moderate | 99 | 57% |
| Severe | 35 | 21% |
| **Number of Prior Surgeries** |  |  |
| 0 | 24 | 14% |
| 1 to 4 | 99 | 56% |
| 5 to 8 | 22 | 13% |
| 9 or more | 29 | 17% |

#### Construct Validity

An explanatory factor analysis was utilized to examine the factorability of the 21 item QUEST measure. The data were screened for univariate outliers, and the minimum amount of data for factor analysis was satisfied. All bivariate correlations between the items were less than .80. Bartlett’s test of sphericity was significant and demonstrated adequacy (χ^2^ = (210) = 2491.83, *p* < .001). Kaiser-Meyer-Olkin measure of sampling adequacy was .927, above the recommended value of .50 (Field, 2013). Diagonals of the anti-image correlation matrix exceeded .3, supporting inclusion of each item in the factor analysis (Cerny & Kaiser, 1977). Communalities ranged between .445 and .784, confirming all other items shared some common variance. Eigenvalues were based on a non-forced identification, allowing the measure to naturally identify the number of factors. Total overall variance explained was 61.81%. The first factor explained 46.83% of the variance, the second factor explained 6.79% of the variance, the third factor explained 4.43% of the variance, and the fourth factor explained 3.76% of the variance (Supplemental Table 2). Factors appear to reflect the holistic importance of psychosocial distress (Factor 1), with additional factors focused on the unique impacts of motor impairments and pain (Factor 2), lower cranial nerve dysfunction (Factor 3), and hearing problems (Factor 4). An Oblimin with Kaiser Normalization was evaluated across the four factors. All but two questions (vision and work/school) loaded above the minimum threshold of .30 (Table 4). Overall, the 21-item model, 4-factor model indicated adequate fit.

**Table 4.** Factor Analysis Rotation.

|  | **Factor** |  |  |  |
| --- | --- | --- | --- | --- |
|  | **1** | **2** | **3** | **4** |
| hearing | -0.017 | 0.161 | 0.080 | **-0.593** |
| pain | 0.263 | **0.402** | 0.080 | 0.051 |
| mobility | -0.083 | **0.817** | 0.050 | -0.116 |
| vision | 0.260 | 0.152 | 0.192 | -0.153 |
| balance | -0.103 | **0.764** | 0.089 | -0.124 |
| facial_weakness | -0.037 | 0.036 | **0.697** | -0.091 |
| eating_drinking | 0.053 | -0.003 | **0.813** | 0.033 |
| speaking | -0.012 | 0.061 | **0.761** | -0.037 |
| fingers_hands | 0.062 | **0.600** | 0.204 | 0.289 |
| adjustments | 0.114 | **0.596** | 0.090 | -0.201 |
| dependent | 0.314 | **0.469** | 0.068 | -0.128 |
| leisure | 0.200 | **0.599** | -0.109 | -0.280 |
| work_school | 0.354 | 0.332 | 0.037 | -0.237 |
| avoid_social | 0.175 | 0.145 | 0.126 | **-0.575** |
| relationships | **0.457** | 0.183 | 0.081 | -0.276 |
| communication | 0.096 | -0.025 | 0.128 | **-0.757** |
| isolated | **0.456** | -0.076 | 0.240 | -0.409 |
| anxious | **0.755** | -0.131 | 0.064 | -0.088 |
| self_conscious | **0.671** | -0.061 | 0.204 | -0.049 |
| disempowered | **0.891** | 0.200 | -0.152 | -0.013 |
| unsure_future | **0.771** | 0.026 | 0.011 | 0.072 |

**Legend:** Item loadings on four factors identified in an exploratory factor analysis (EFA) using an oblique (Oblimin) rotation with Kaiser normalization. Values indicate the strength and direction of the association between each item and factor; loadings $\geq 0.40$ or $\leq -0.40$ are shown in bold.

#### Internal Reliability and Test/Retest Reliability

Internal reliability of QUEST was excellent, with a Cronbach’s alpha of 0.946. The test-retest reliability of the scale was assessed using data from 81 participants invited to complete the measure a second time. Amongst these 81 participants, the mean QUEST total score was 55.40 (SD=18.97) on first administration and 53.78 (SD=18.61) on second administration. Cronbach’s Alpha was 0.952 across both administrations, and the inter-item correlation coefficient between administrations was .909, further supporting the reliability of the scale and confirming its reliability over repeated measurements. The ANOVA results showed no significant difference between items (F = 3.283, p = .074), providing additional evidence that the measurements were consistent over time.

#### Convergent Validity

In establishing convergent validity, bivariate correlations across the Global Severity Scale, NFTI-QoL, SAC, FACT-G, PROMIS Pain Interference, PROMIS Cognitive Function, were evaluated to that of the QUEST, as seen in Table 5. There were significant correlations in the expected directions with the global severity scale (.824), NFTIQOL (.856), PROMIS CF (-.583), SAC (.541), PROMIS PI (.479), and the FACT-G (-.429). Similar correlations were noted between these measures and the NFTI-QoL, another measure of *NF2*-related quality of life.

**Table 5.** Bivariate Correlations.

|  | **QUEST** | **Global**<br>**Severity** | **PROMIS**<br>**CF** | **NFTIQOL** | **SAC** | **FACT-G** | **PROMIS**<br>**PI** |
| --- | --- | --- | --- | --- | --- | --- | --- |
| **QUEST** |  | .824** | -.583** | .856** | .541** | -.429** | .479** |
|  |  | 0.000 | 0.000 | 0.000 | 0.000 | 0.000 | 0.000 |
| **Global**<br>**Severity** | .824** |  | -.430** | .736** | .488** | -.254** | .466** |
|  | 0.000 |  | 0.000 | 0.000 | 0.000 | 0.001 | 0.000 |
| **PROMIS**<br>**CF** | -.583** | -.430** |  | -.609** | -.328** | .366** | -.431** |
|  | 0.000 | 0.000 |  | 0.000 | 0.000 | 0.000 | 0.000 |
| **NFTIQOL** | .856** | .736** | -.609** |  | .475** | -.333** | .563** |
|  | 0.000 | 0.000 | 0.000 |  | 0.000 | 0.000 | 0.000 |
| **SAC** | .541** | .488** | -.328** | .475** |  | -.188* | .173* |
|  | 0.000 | 0.000 | 0.000 | 0.000 |  | 0.015 | 0.025 |
| **FACT-G** | -.429** | -.254** | .366** | -.333** | -.188* |  | -0.093 |
|  | 0.000 | 0.001 | 0.000 | 0.000 | 0.015 |  | 0.230 |
| **PROMIS**<br>**PI** | .479** | .466** | -.431** | .563** | .173* | -0.093 |  |
|  | 0.000 | 0.000 | 0.000 | 0.000 | 0.025 | 0.230 |  |

\*\*Correlation is significant at the 0.01 level (2-tailed)
\* Correlation is significant at the 0.05 level (2-tailed)

#### Incremental Validity

Evaluation of the incremental validity came from hierarchical linear regression. As noted in Table 6, the NFTIQOL, SAC, FACT-G, and PROMIS PI scales were sequentially entered into the model. NFTIQOL accounted for the previously standardized *NF2*-specific QoL, SAC accounted for hearing function and hearing-related quality of life, FACT-G evaluates overall well-being and generic quality of life, and PROMIS PI accounted for pain interference. Significant differences were noted through Step 1 through 3, and for most of Step 4 (except PROMIS PI). QUEST was a significant predictor of the behaviors (disease-specific QoL, hearing function, and overall wellbeing), all critical for individuals with *NF2*-SWN.

**Table 6.** Hierarchical Linear Regression.

|  | **Variable** | **B** | **Adjusted**<br>**$R^2$** | **SE** | **$\beta$** | **t** | **P** | **CI**<br>**Lower**<br>**Bound** | **CI**<br>**Upper**<br>**Bound** |
| --- | --- | --- | --- | --- | --- | --- | --- | --- | --- |
| Step 1 | NFTIQOL_TOTAL | 3.903 | .724 | 0.186 | 0.852 | 20.950 | 0.000 | 3.535 | 4.271 |
| Step 2 | NFTIQOL_TOTAL | 3.520 | .746 | 0.203 | 0.768 | 17.345 | 0.000 | 3.119 | 3.921 |
|  | SAC | 0.338 |  | 0.085 | 0.176 | 3.965 | 0.000 | 0.170 | 0.507 |
| Step 3 | NFTIQOL_TOTAL | 3.264 | .767 | 0.205 | 0.712 | 15.949 | 0.000 | 2.860 | 3.668 |
|  | SAC | 0.332 |  | 0.082 | 0.172 | 4.059 | 0.000 | 0.170 | 0.493 |
|  | FACT_G | -0.319 |  | 0.080 | -0.160 | -3.989 | 0.000 | -0.477 | -0.161 |
| Step 4 | NFTIQOL_TOTAL | 3.112 | .768 | 0.248 | 0.679 | 12.561 | 0.000 | 2.623 | 3.601 |
|  | SAC | 0.343 |  | 0.082 | 0.178 | 4.167 | 0.000 | 0.181 | 0.506 |
|  | FACT_G | -0.332 |  | 0.081 | -0.166 | -4.105 | 0.000 | -0.491 | -0.172 |
|  | PROMIS_Pain Interference | 0.103 |  | 0.095 | 0.050 | 1.089 | 0.278 | -0.084 | 0.290 |

a. Dependent Variable: QUEST\_Total

#### Known-Groups Validity

Known-groups validity of the measure was demonstrated as the measure significantly distinguished patients based on self-reported disease severity (ANOVA F-value 50.788, P<0.001) and number of prior *NF2*-related surgeries undergone (ANOVA F-Value 13.953, P<0.001) (Table 7).

**Table 7.** QUEST Score for Total Sample and Stratified by Self-Reported Disease Severity and Number of Prior *NF2*-Related Surgeries.

|  | **N** | **Mean** | **SD** | **Minimum** | **Maximum** |
| --- | --- | --- | --- | --- | --- |
| **Total** | 174 | 57.33 | 19.96 | 21 | 103 |
| **Disease Severity** |  |  |  |  |  |
| **No Symptoms** | 3 | 24.33 | 5.77 | 21 | 31 |
| **Mild** | 36 | 37.19 | 13.03 | 23 | 71 |
| **Moderate** | 99 | 58.33 | 15.42 | 28 | 94 |
| **Severe** | 36 | 77.47 | 14.18 | 45 | 103 |
| **Number of *NF2* surgeries** |  |  |  |  |  |
| **0** | 24 | 46.13 | 20.59 | 21 | 85 |
| **1-4** | 99 | 53.24 | 17.41 | 23 | 87 |
| **5-8** | 22 | 71.50 | 16.34 | 38 | 94 |
| **9 or more** | 29 | 69.83 | 19.12 | 38 | 103 |

Legend: SD = standard deviation

## Discussion

The present study used a three-phase, mixed methods approach to develop and examine the content validity and initial psychometric properties of the QUEST. Analyses supported the validity of a 21-item measure that describes the diverse impacts of *NF2*-SWN symptoms on participant’s day-to-day lives. The measure showed robust content validity based on input from patient and clinician experts. Initial testing of the measure in an international cross-sectional cohort showed good internal consistency and test-retest reliability, with evidence for convergent, incremental, and known-groups validity of the total QUEST score.

The QUEST assessment represents an important advance in measuring disease-specific QoL for individuals living with *NF2*-SWN. As these individuals typically live with multiple tumor types across multiple body locations, as well as other non-tumor related symptoms, measuring disease-specific quality of life allows for targeted yet comprehensive assessment of patients’ top concerns not achievable with generic quality of life measures. Disease-specific measures like the QUEST and NFTI-QOL are thus important tools to track the impact of *NF2*-SWN and its treatments on patients’ QoL across disease stages (e.g. in diseases registries or natural history studies). Initial validation of the QUEST adds an important option for disease-specific measures for NF2-SWN; clinicians and researchers who require a shorter measure, longitudinal comparative data, or validated translations may prefer the NFTI-QOL, while those seeking a more detailed assessment of symptoms or who are aiming to assess the impact of new treatments on *NF2*-SWN patients’ quality of life may may prefer the QUEST. As the QUEST includes physical, emotional, and social items, it is likely to be useful in measuring the impact of the wide array of treatments employed for this complex disease, including drugs, devices, surgical approaches, radiation, psychological interventions and behavioral therapies.

The QUEST assessment is further supported as a clinically meaningful outcome measure, as the pattern of findings from hierarchical modeling supports the QUEST total score aligns with how patients function and feel in day-to-day life. The QUEST total score tracked closely with disease-specific quality of life, and reflected hearing-related functioning—an essential domain in *NF2*-SWN that often shapes communication, participation, and overall independence. Concurrently, the QUEST total score was moderately associated with broader perceptions of general well-being, suggesting that it complements—rather than duplicates—existing quality-of-life measures by capturing patient-centered impacts that reflect everyday functioning and lived experience with NF2-SWN. The limited additional contribution of pain interference once other domains were considered may indicate that, for many patients, pain-related disruption is either less central to the overall burden reflected by QUEST or is already embedded within other aspects of disease impact that the QUEST captures.

Data shown in Table 7 further strengthens the clinical relevance of QUEST by showing that the measure distinguishes between groups in ways that mirror clinical expectations and lived experience. Individuals who perceived their disease as more severe and those with a larger history of *NF2*-SWN-related surgeries reported higher QUEST scores, consistent with the notion that accumulated treatment complexity and advanced disease course are reflected in patient-reported burden. Together, these results position QUEST as a practical measure for clinical care and research that is sensitive to meaningful differences in disease impact, relevant to core functional domains (especially hearing), and anchored to clinical milestones that often mark progression or greater complexity. As such, the QUEST score may be well-suited for tracking change in quality of life over time and provide an interpretable endpoint for evaluating interventions in *NF2-*SWN.

Historically, clinical trials for *NF2*-SWN have focused on demonstrating the impact of treatment on tumor volume, with functional assessments limited to hearing performance and limited use of patient-reported outcomes (Chiranth et al., 2023; Plotkin et al., 2009). However, measuring disease-specific QoL is important for the clinical outcome assessment strategy of new treatments for the histologically benign tumors of *NF2*-SWN, where improvement to lifespan is not expected and thus other evidence of clinical benefit to patients is required by the FDA and other regulators (Merker et al., 2024). By using a disease-specific measure like QUEST that has been initially validated according to FDA guidelines, clinical trialists may be better poised to demonstrate whether new treatments *NF2*-SWN benefit patients’ quality of life during regulatory review (Food and Drug Administration, 2009). Furthermore, use of holistic, disease-specific measures in place of narrower tumor- or symptom-specific measures could be especially useful for basket trials testing the effect of interventions across multiple tumor type simultaneously (Plotkin et al., 2024) and trials of gene therapy which may have diverse impacts on treatment and prevention across body systems (Staedtke et al., 2023).

### Clinical Implications

Clinicians could integrate QUEST into routine assessment and care planning to improve early detection of quality of life issues and more precisely individualize treatment. Where results indicate measurable clinical signals, incorporating the QUEST into structured screening, triage, and monitoring workflows can help prioritize patients most likely to benefit from timely intervention, escalation of care, or targeted referrals. In practical terms, this aligns well with a stepped-care approach in which treatment intensity and modality are matched to patient need, supported by consistent measures to track response over time and guide shared decision-making. To sustain implementation, clinical deployment should be paired with ongoing quality improvement that includes auditing adherence to recommended assessment protocols, monitoring patient outcomes longitudinally, and refining clinical treatment and referral pathways as new evidence on this rare disorder emerges.

### Constrains on Generality

Much of our sample was White and female, reflecting the demographic groups in the underlying clinical trial, specialty clinic, and disease registry from which we recruited. While these venues were critical for connecting with the large number of patients with this rare disease needed for rigorous psychometric analysis, outreach to and testing of this measure in more diverse populations is needed. In addition, Phase I and II patient data was collected solely from U.S. residents; Phase I clinician data and Phase III patient data were expanded to include residents of U.S., Canada, the United Kingdom, and Australia but still represent only WEIRD (Western, educated, industrialized, rich, and democratic) countries. As such, study findings may not generalize to the broader global population. Future research is needed to confirm the cultural relevance of items in more diverse cohorts, and validate the survey in additional languages and settings.

### Additional Limitations and Future Directions

The results of this study should be interpreted considering several additional limitations. While we demonstrated multiple aspects of the construct validity of the QUEST, we did not evaluate discriminant validity nor assess the relationship between our measure and objective disease indices or clinician-reported disease severity. Future research could examine how QUEST scores relate to standardized performance metrics reflective of the identified factors, including tests of hearing, facial function, and muscle weakness (Akshintala et al., 2021; Plotkin et al., 2013). Additionally, our initial validation survey was cross-sectional; future work must incorporate the assessment into longitudinal studies to determine whether it is responsive to change over time and suitable for assessing treatment impact in clinical trials. Finally, our measure was validated only for individuals ages 15+; while most *NF2*-SWN patients are diagnosed after this age (Forde et al., 2021), future research may be performed to develop a version appropriate for younger adolescents, who may have unique symptom concerns (Ruggieri et al., 2016) and/or require simpler item wording.

### Conclusions

Despite these limitations, the present study presents a robustly developed measure of *NF2*-SWN quality of life that addresses limitations in prior disease-specific patient-reported measures and expands the clinical outcome assessment toolbox for *NF2*-SWN clinical trials. While the NFTI-QoL will remain a valuable assessment tool, particularly in routine clinical care where rapid assessment and comparison to established norms is a priority, the QUEST can fill a specific niche in clinical trial design for *NF2*-SWN. Importantly, we believe this measure will have improved content validity in clinical trial participants ages ≥15 years and improved sensitivity to change over the typical 1-2 year timeline of clinical trials when compared to existing measures. It also prioritizes feasibility in trial settings by being free, short, and easy to administer and score online. The QUEST has already been added to the study protocols for two upcoming *NF2*-SWN clinical trials, which will provide necessary validation data on the measure’s sensitivity to change and ability to assess patients’ responses to new treatments. Data generated by the QUEST can then be considered by clinicians, researchers, and regulatory agencies like the FDA to provide additional evidence of the potential clinical benefit of new treatments for *NF2*-SWN.

## Data Availability

Data for Phase I of the research is not publicly available; data for Phase II and Phase III will be publicly available no later than October 1, 2027 at the NF Data Portal and can be accessed at https://nf.synapse.org/Explore/Studies/syn53181385/. Analysis code is not available for the study; however, other research materials are available on request from the first author.

## Acknowledgements

The authors thank the investigators and staff of the INTUiTT-NF2 clinical trial for recruiting and consenting participants for Phase I of the study. The authors also acknowledge the entities that provided funding for the brigatinib arm of the INTUITT-NF2 trial in which our qualitative sub-study was embedded; this includes the Children’s Tumor Foundation, Takeda Pharmaceuticals, the Andrea Cahill Foundation, and the National Cancer Institute (Grant P30CA006516). The authors thank the Response Evaluation in Neurofibromatosis and Schwannomatosis (REiNS) International Collaboration Patient Reported Outcomes working group for their feedback on the draft QUEST measure during Phase II of the study. Finally, we would like to thank the many NF non-profit organizations who assisted in recruiting for Phase III of the study, which includes the Children’s Tumor Foundation (U.S.), Children’s Tumour Foundation (Australia), NF Network, Nerve Tumours UK, NF2 Biosolutions, NF Northeast, NF Midwest, Texas NF, NF North Central, NF Arizona/Southwest, NF Michigan, NF Central Plains, NF California, and NF Tennessee.

## Conflicts of Interest

FB is a consultant for Addinex Technologies (New York, NY). SP is a co-founder of NF2 Therapeutics (Cambridge, MA). VM has received in-kind research support from Captify, Inc. (San Francisco, CA). The remaining authors have no relevant conflicts of interest to disclose.

## Funding

This study was funded by Children’s Tumor Foundation (CTF) Clinical Research Awards 2020-10-001 and 2023-10-003 to VM.

---

**Supplemental Table 1.**
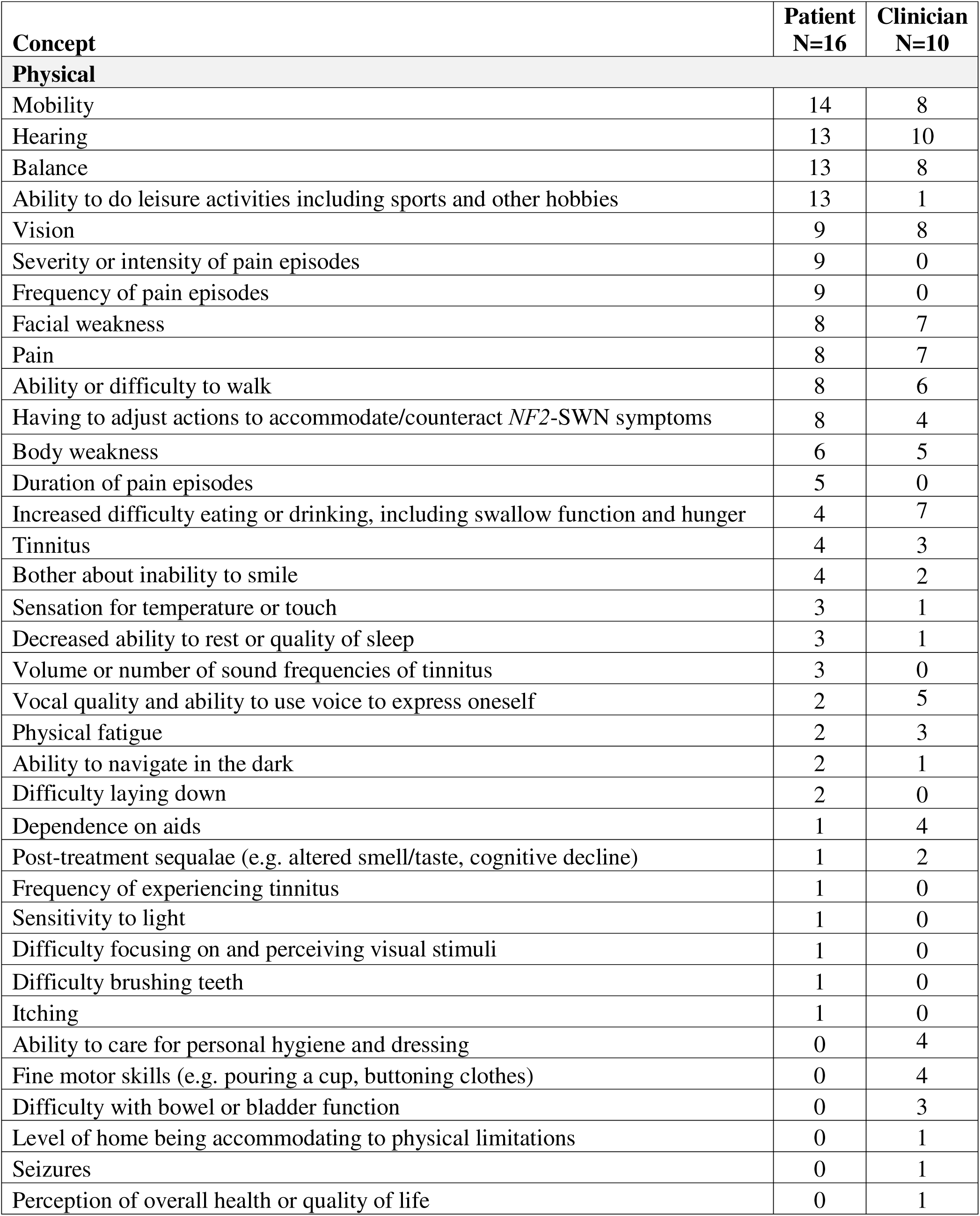
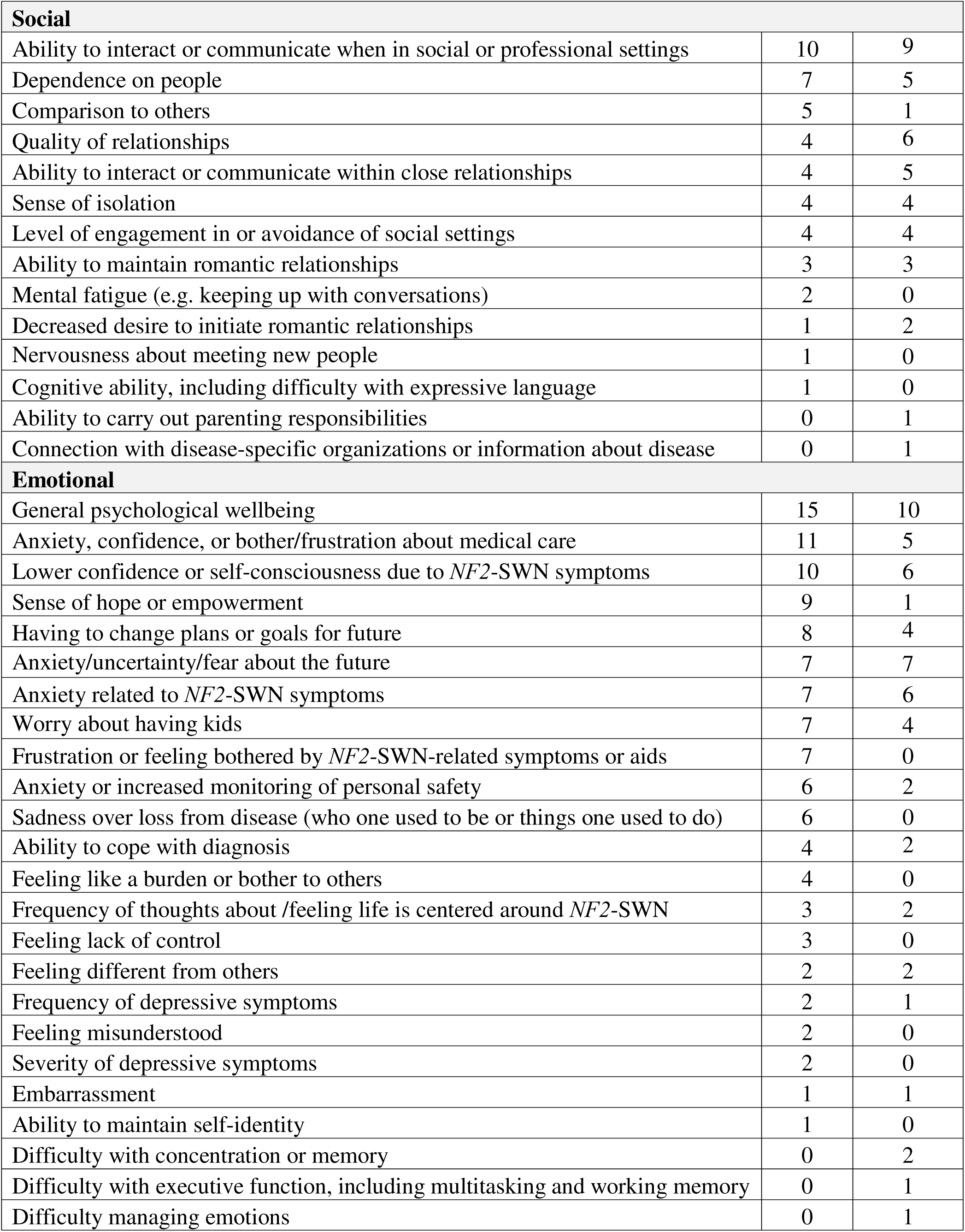
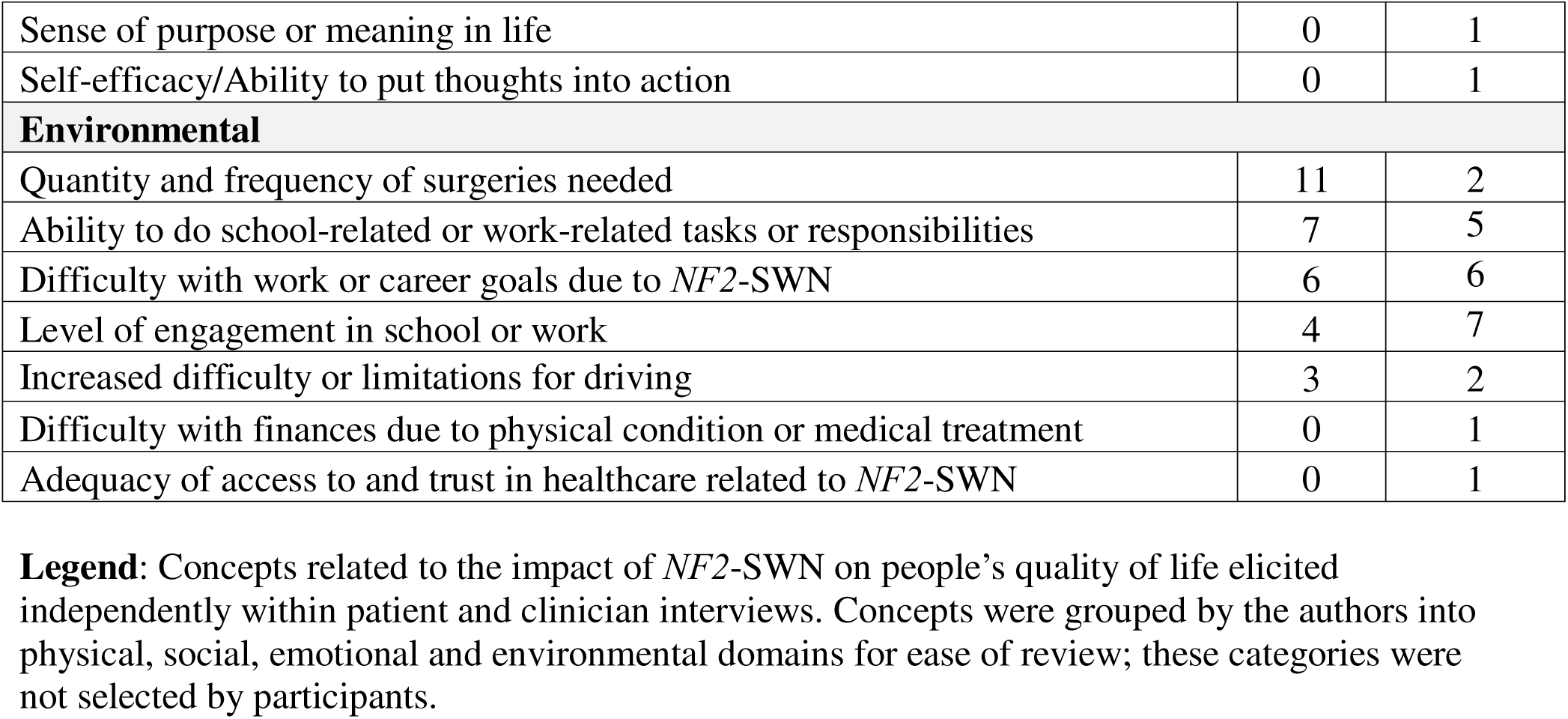
Concepts Elicited in Patient and Clinician Interviews.

**Supplemental Table 2.**
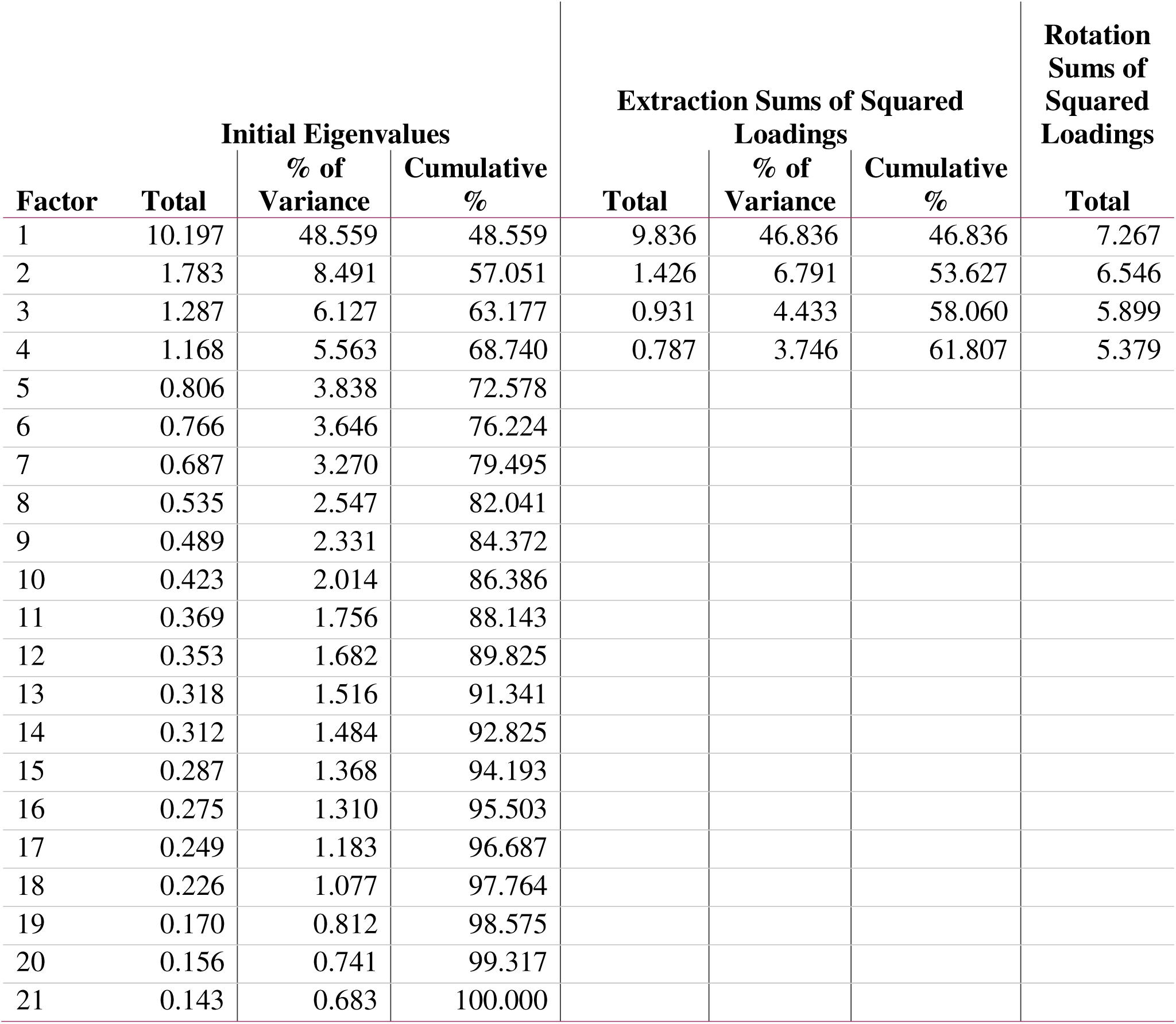
QUEST Factor Analysis.

## Notes

### Author Declarations

All study procedures were reviewed and approved by the Dana Farber/Harvard Cancer Center Institutional Review Board (Protocol 19-828) or the Mass General Brigham Institutional Review Board (Protocol 2023P003305 and Protocol 2024P003196).

