## Supplemental Appendix A for "Development and Initial Validation of the Quality of life Evaluation in *NF2*-related Schwannomatosis Trials (QUEST) Assessment"

**<u>Qu</u>ality of life <u>E</u>valuation in *NF2*-related <u>S</u>chwannomatosis <u>T</u>rials (QUEST)**

Please note, NF2-related schwannomatosis (formerly called neurofibromatosis type 2) will be abbreviated as “NF2” in the following questions.

These questions will ask you about how much NF2 symptoms have impacted you **in the past <u>7 days</u>**. You may not have all of these symptoms. The scale goes from <u>not at all</u> to <u>very much</u>.

Please select one response per question and be as honest as possible.

| **In the past <u>7 days</u>…** | Not at All<br>1 | 2 | Somewhat<br>3 | 4 | Very Much<br>5 |
|---|---|---|---|---|---|
| 1. How much has NF2-related hearing loss interfered with your day-to-day activities (taking into account any hearing devices if you use them)? | ☐ | ☐ | ☐ | ☐ | ☐ |
| 2. How much has NF2-related pain interfered with your day-to-day activities? | ☐ | ☐ | ☐ | ☐ | ☐ |
| 3. How much have NF2-related mobility issues interfered with your day-to-day activities? | ☐ | ☐ | ☐ | ☐ | ☐ |
| 4. How much have NF2-related vision issues interfered with your day-to-day activities? | ☐ | ☐ | ☐ | ☐ | ☐ |
| 5. How much have NF2-related balance issues interfered with your day-to-day activities? | ☐ | ☐ | ☐ | ☐ | ☐ |
| 6. How much has NF2-related facial weakness impacted your day-to-day functioning? | ☐ | ☐ | ☐ | ☐ | ☐ |
| 7. How much have NF2-related symptoms interfered with your eating or drinking? | ☐ | ☐ | ☐ | ☐ | ☐ |
| 8. How much have NF2-related symptoms interfered with your speaking? | ☐ | ☐ | ☐ | ☐ | ☐ |
| 9. How much have NF2-related symptoms interfered with using your fingers and hands in your day-to-day activities? | ☐ | ☐ | ☐ | ☐ | ☐ |

| In the past <u>7 days</u>… | Not at All 1 | 2 | Somewhat 3 | 4 | Very Much 5 |
|---|---|---|---|---|---|
| 10. How much have NF2-related symptoms required you to make adjustments in your day-to-day activities? | ☐ | ☐ | ☐ | ☐ | ☐ |
| 11. How much have NF2-related symptoms made you feel dependent on others? | ☐ | ☐ | ☐ | ☐ | ☐ |
| 12. How much have NF2-related symptoms interfered with your ability to participate in your current hobbies, sports, or leisure activities? | ☐ | ☐ | ☐ | ☐ | ☐ |
| 13. How much have NF2-related symptoms interfered with doing your tasks at work or school? | ☐ | ☐ | ☐ | ☐ | ☐ |
| 14. How much have NF2-related symptoms made you avoid social events? | ☐ | ☐ | ☐ | ☐ | ☐ |
| 15. How much have NF2-related symptoms impacted your relationships with family and friends? | ☐ | ☐ | ☐ | ☐ | ☐ |
| 16. How much have NF2-related symptoms affected your communication with others? | ☐ | ☐ | ☐ | ☐ | ☐ |
| 17. How much have NF2-related symptoms made you feel socially isolated? | ☐ | ☐ | ☐ | ☐ | ☐ |
| 18. How much have NF2-related symptoms made you feel anxious? | ☐ | ☐ | ☐ | ☐ | ☐ |
| 19. How much have NF2-related symptoms made you feel self-conscious about what other people might think of you? | ☐ | ☐ | ☐ | ☐ | ☐ |
| 20. How much have NF2-related symptoms made you feel disempowered? | ☐ | ☐ | ☐ | ☐ | ☐ |
| 21. How much have NF2-related symptoms made you feel unsure about your future? | ☐ | ☐ | ☐ | ☐ | ☐ |

**This last question asks about your NF2-related symptoms overall.**

22. How much have NF2-related symptoms interfered with your day-to-day activities in the past 7 days?

| Not at all | | | | | Somewhat | | | | | Very Much |
|---|---|---|---|---|---|---|---|---|---|---|
| 0 | 1 | 2 | 3 | 4 | 5 | 6 | 7 | 8 | 9 | 10 |
| ☐ | ☐ | ☐ | ☐ | ☐ | ☐ | ☐ | ☐ | ☐ | ☐ | ☐ |
